# Local field potential signal transmission is correlated with the fractional anisotropy measured by diffusion tractography

**DOI:** 10.1101/2024.04.14.24305803

**Authors:** Maral Kasiri, Sumiko Abe, Rahil Soroushmojdehi, Estefania Hernandez-Martin, Alireza Seyyed Mousavi, Terence D. Sanger

## Abstract

**Objective:** In this paper we aim to examine the correlation between diffusion tensor imaging (DTI) parameters of anatomical connectivity and characteristics of signal transmission obtained from patient-specific transfer function models. Here, we focused on elucidating the correlation between structural and functional neural connectivity within a cohort of patients diagnosed with dystonia.

**Methods:** DTI images were obtained from twelve patients with dystonia prior to the deep brain stimulation (DBS) surgery. For each patient we processed the imaging data to estimate anatomical measures including fractional anisotropy (FA), axial diffusivity (AD), number of fiber tracts per unit area (N), and fiber tract length (L). After the implantation of temporary depth leads for each patient as part of their treatment plan, intracranial signals were recorded. Transfer function models and the corresponding measures of functional connectivity were computed for each patient using local field potential (LFP) recordings. Linear mixed effect analysis was then employed to determine the relationship between transfer function measures and DTI parameters.

**Results:** Our results illustrate a positive correlation between FA, AD, and intrinsic neural transmission measures, representing amplification and spread of intrinsic neural oscillations, obtained from the transfer functions models. However, no significant correlation was found between the functional connectivity and number of fiber tracts or fiber lengths.

**Conclusion:** Our findings suggest that white matter integrity, as measured by FA and AD, can potentially reflect the amplification and spread of intrinsic brain signals throughout the network. This study underscores the significant relationship between structural and functional connectivity, offering valuable insights into propagation of neural activity in the brain network and potential implications for optimizing noninvasive treatments and planning for neurological disorders.

## I. Introduction

Advancements in neuroimaging techniques have fundamentally changed our understanding of brain functional and anatomical connectivity [1], [2]. Diffusion Tensor Imaging (DTI), as an advanced magnetic resonance imaging (MRI) modality, enables us to visualize white matter tracts that connect cortical and subcortical structures by measuring the motion of water molecules and provide us with valuable insights into the structural connectivity of the brain. With its capacity to reveal complex details of brain micro-structure, DTI plays a significant role in optimizing procedures like deep brain stimulation (DBS) [3]–[12]. DBS procedure can be finely tuned by utilizing DTI to precisely map neural pathways and understand microstructural connections, promising improved treatment protocols and outcomes for patients suffering from neurological disorders [5].

Studying the relationship between structural and functional connectivity is an important domain of research for understanding the brain as a complex network of interconnected regions [13]–[17]. Previous studies have explored the complex relationships and communication patterns between different brain areas in various neurological disorders. For example such analysis in epilepsy helps to map the seizure network by exploring the relationships between the structural and functional networks responsible for conduction of epileptic activity [16]. Moreover, such studies have been conducted on healthy subjects to find the correlation between DTI measures and resting state functional MRI (fMRI). fMRI has lower spatial resolution than electrophysiology data and it does not capture the deep brain activity. On the other hand, it clearly reveals the cortical functional connectivity. This study showed that functional connectivity reflects structural connectivity to a large degree in cortical regions, although there is not a definite one-to-one mapping [18]. Despite all the research endeavors that provide invaluable insights into the interaction between structural and functional connectivity, the relationship between the two for transmission of non-epileptic brain signals remains unclear.

Transfer functions have been widely used to study the signal transmission and physiological connectivity within brain regions [19], [20]. For example, Kamali et al. [20] utilized characteristics of patient specific transfer function models of pathways in order to localize the seizure onset zones. In our previous studies, we have shown that transfer function models of deep brain regions can be used to replicate the evoked responses from DBS, informing us of existence of some relationship between the transmission of DBS pulses through neural pathways and the functional connectivity [21]. Moreover, we have shown that the DBS changes the transfer function gains of deep brain regions meaning that DBS pulses have some effects on the functional connectivity. Building upon these fundamental studies that use transfer function methods, we aim to compute measures of signal transmission using patient specific transfer function models and find their relationship with structural characteristics provided by DTI measures, including fractional anisotropy (FA), axial diffusivity (AD), fiber length (L), and number of fibers per unit area (N), in basal ganglia and thalamic subnuclei. We hypothesize that the measures of signal amplification and transmission computed from transfer functions are positively correlated with the FA and AD. In other words, we hypothesise that the white matter integrity of the fibers and the diffusivity of neural pathways are reflected in functional connectivity represented by the transfer function measures.

In order to do so, we recorded signals from deep brain regions in twelve patients with dystonia, who underwent DBS procedure as part of their clinical evaluation. MRI, CT and DTI images were acquired and the DTI anatomical measures were calculated. Transfer functions representing each pathway for all patients were computed. We then compared the DTI measures and the transfer function signal transmission quantities using linear mixed effect model. Understanding the correlation and mapping between DTI parameters that can be calculated noninvasively and characteristics of transfer function models, which require invasive measurements from deep brain regions, offers invaluable insights into the relationship of brain structure and function and results in improvement of treatment protocols, planning, and outcomes.

## II. Materials and Methods

### A. Subjects

twelve pediatric and young adults patients, diagnosed with primary or secondary dystonia by a movement disorder specialist (T.D.S), underwent a staged implantation of DBS leads a [22], [23]. The patients or the guardians of minors provided Health Insurance Portability and Accountability Act (HIPAA) authorization for research use of electrophysiological data before the procedure and provided written informed consent for surgical procedures conforming to standard hospital practice at Children’s Health Orange County (CHOC) or Children’s Hospital Los Angeles (CHLA). The research use of data was approved by the institutional review board of the Children’s Health, Orange County (CHOC) hospital or Children’s Hospital Los Angeles (CHLA). Table.I includes demographic information of all patients participated in this study.

**TABLE 1:** Patients Demographics, including 3 female and 9 male subjects, ages between 5-21 year-old. CP: Cerebral Palsy, GA1: Glutaric aciduria type 1 [24], KMT2B: Lysine Methyltransferase-2B [25]; VA: ventral anterior; VoaVop: ventral oralis anterior-posterior; VIM: ventral intermedia; VPL: ventral posterolateral; STN: subthalamic nucleus; GPi: globus pallidus internus; PPN: pedunculopontine nucleus.

| Subject | Etiology | Implanted Regions |
| --- | --- | --- |
| S1 | CP | VA, VoaVop, VIM, GPi |
| S2 | Unknown (dx CP) | VA, VoaVop, VIM, GPi |
| S3 | CP | VoaVop, VIM, VPL, STN, GPi |
| S4 | CP | VA, VoaVop, VIM, GPi |
| S5 | H-ABC (genetic) | VoaVop, VIM, STN, GPi |
| S6 | Dyskinetic CP | VA, VoaVop, VIM, STN, GPi |
| S7 | GA1 | VA, VoaVop, VIM, STN, GPi |
| S8 | KMT2B | VA, VoaVop, VIM, PPN, STN, GPi |
| S9 | GA1 | VA, VoaVop, VIM, PPN, STN, GPi |
| S10 | CP | VA, VoaVop, VIM, PPN, STN, SNr, GPi |
| S11 | MYH2 | VoaVop, VIM, PPN, STN, SNr, GPi |
| S12 | Dyskinetic CP | VoaVop, VIM, PPN, STN, SNr, GPi |

### B. Surgical Procedure and Depth Recording

For each patient, up to 12 temporary AdTech MM16C depth sEEG leads (AdTech Medical Instrument Corp., Oak Creek, WI, United States) were implanted under general anesthesia in potential DBS targets depending on diagnosis and clinical symptoms using standard stereotactic procedure [22], [23]. Potential stimulation targets were identified based on previous studies of clinical efficacy. These targets include subthalamic nucleus (STN), globus pallidus internus (GPi), in basal ganglia, ventral intermediate nucleus (VIM), ventral oralis anterior/posterior (VoaVop), and ventral anterior nucleus (VA), and ventral postrolateral nucleus (VPL), in thalamus, pedunculopontine nucleus (PPN) and Substantia Nigra reticulata (SNr) in the brainstem [26], [27].

The implanted sEEG leads contain ten high-impedance (70–90 kΩ) micro-wire electrodes (50-µm diameter) that are arranged in groups of two or three, spaced evenly around the circumference of the lead shaft in four rows. The leads were connected to Adtech Cabrio™ connectors that include a custom unity-gain preamplifier for each micro-contact to reduce noise and motion artifacts. All data used in this study were recorded through the high impedance contacts, with stimulation off, and sampled at 24 kHz by a custom recording system consisting of a PZ5M 256-channel digitizer, RZ2 processor, and RS4 high speed data storage (TDT, Tucker-Davis Technologies Inc., Alachua, FL, United States).

### C. Structural Connectivity

#### 1) Structural image acquisition and processing

Preoperative MRI T1-weighted (structural MRI), DTI, and postoperative CT scans were obtained. Subsequently, the following steps were taken: 1) Pre-processing to correct distortions and motion artifacts of the DTI scans based on TOPUP and Eddy Current Correction algorithms [28]; 2) Co-registration of the DTI and postoperative CT images to the T1 anatomic volume; 3) Segmentation of all the subregions of the thalamus and pallidum; 4) Localization of sEEG leads using the attenuation in CT images; and 5) Estimation of micro-contacts’ coordinates based on a linear model of the sEEG lead and assigning 3 mm diameter to the effective area [29].

#### 2) Fiber tracking and DTI parameter estimation

In our study, DTI was employed to explore fiber orientations and measure diffusion properties. Micro-contact regions within each nucleus served as both origin and target regions for fiber tracking, enabling visualization of the anatomic pathways connecting the origin and the target. Using the diffusion tensor we computed several metrics to further characterize the fiber tracts linking these regions: 1) AD, calculated as the first eigenvalue (*λ*_1_) of the diffusion tensor, reflects water molecule diffusion along the principal axis of fiber tracts, indicating axonal integrity. Changes in AD can indicate axonal degeneration or demyelination [30]. 2) FA, a measure derived from the variance of eigenvalues (*λ*_1_, *λ*_2_, *λ*_3_), quantifies the degree of anisotropy in water diffusion, offering insights into the coherence and density of fiber tracts. The calculation of FA is given by Equation 1, which is scaled between 0 (isotropic diffusion) and 1 (highly anisotropic diffusion). This provides direct insight into the structural integrity of axonal fibers [31]. 3) Number of fiber tracts per unit area (N) estimates the count of individual fiber bundles connecting two regions of interest and offers information about the density of fibers within that area [32]. 4) Fiber tract length indicates the total length of individual fiber tracts connecting two regions of interest and provides insights into the spatial extent or reach of neural pathways [33].

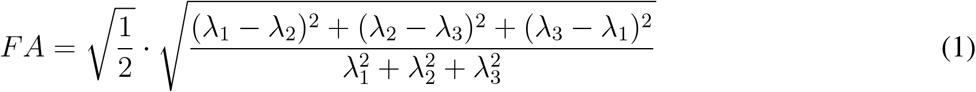

### D. Functional Connectivity

#### 1) Electrophisological data processing

The LFP recordings from 10 micro-contacts of each lead were notch filtered at 60 Hz and its 5 harmonics. They were then high pass filtered at 1 Hz to remove the drift. Each adjacent pair of micro-contacts recordings were subtracted from each other to capture their voltage difference (bipolar montage), which essentially removes the common noise and reveals the underlying neural activity.

#### 2) Transfer Function Computation

The empirical transfer function of a system is computed as the ratio of the system’s output Fourier transform (FT(Y)) to the system’s input Fourier transform (FT(X)) which can be estimated as:

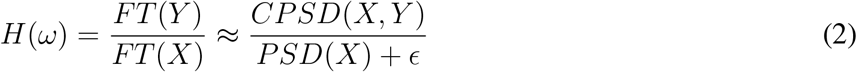

Where PSD(X) is the power spectral density of the input, CPSD(X,Y) is the cross power spectral density between input signal and output signal, with complex numbers, and *ϵ* is a regularization constant. We computed the single input-single output (SISO) transfer function model between each two bipolar recording channels. After constructing the SISO transfer function models for each pair of channels, we want to investigate whether properties of magnitude of these transfer functions correlate with the anatomical features derived from DTI data. In other words, we want to evaluate how the characteristics of these transfer functions, representing functional connectivity, are related to the anatomical connectivity in the brain and to confirm whether they contain useful information about the anatomical features of the neural fibers. Note that the computed transfer functions are complex functions, which includes information about the phase and magnitude. Here, we only use the magnitude of these transfer functions, and not the phase shifts and delays [21].

We computed two parameters representing the characteristics of intrinsic signal transmission from transfer function models as depicted in Figure 1. First parameter is the peak gain or the maximum transfer function gain (*P*_1_ in Figure 1), which represents the maximum level of amplification of the transmitted input signal. In other words, it is a metric that quantifies how much input signals can be amplified and spread in the network at the maximum gain frequency. The larger the gain, the more propagation and amplification of neural activity throughout the network at that frequency. The second parameter is the peak-to-floor (PF) ratio which represents the large system responses and its fast magnitude drop-off [20]. PF ratio is calculated as the ratio between peak of the frequency response and its magnitude at the roll-off frequency (*P*_2_ in Figure.1). The roll-off frequency is the boundary where the energy flowing through a system begins to drop, defined as the frequency at which the dB magnitude is 3dB below the gain at frequency 0 *ω* = 0 (DC gain) or where the power drops to half the power at *ω* = 0, as *P*_2_ in Fig.1. For a given pathway, we calculated the frequency response magnitude of all the SISO transfer functions as quantified by the magnitude of *H*(*ω*). Thereafter, we computed the PF ratio as:

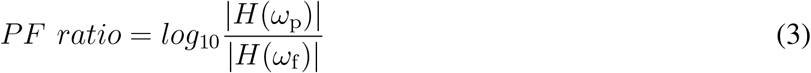

where *ω*_p_ represents the frequency at which maximum gain (*H*(*ω*_p_)) is achieved and *ω*_f_ is roll-off frequency and *H*(*ω*_f_) is the transfer function gain at the roll-off frequency. All these measure were computed for each pair of electrodes, per hemisphere.

**Fig. 1:**
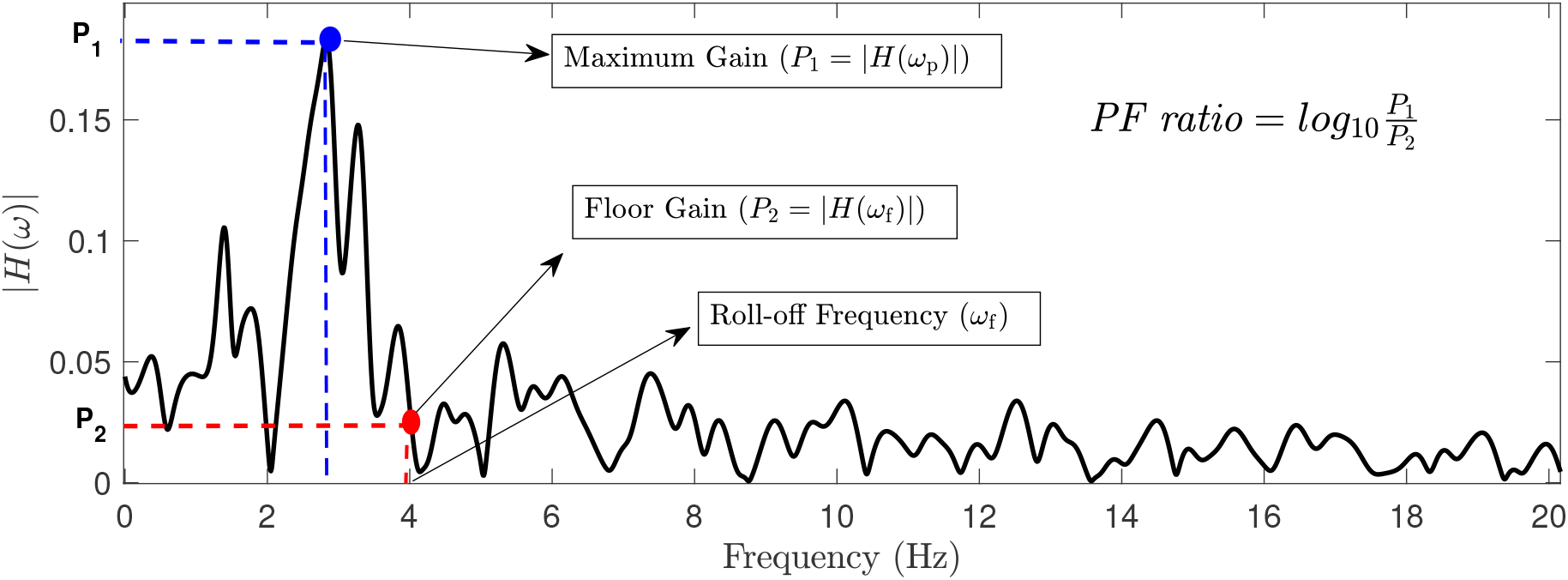
This figure illustrates the magnitude plot of a sample transfer function, showcasing key parameters including maximum gain, floor gain, and roll-off frequency. The PF ratio is determined by calculating the ratio between *P*_1_ and *P*_2_.

In our analysis, we focused exclusively on fibers projecting from the GPi to other regions, because of its established role in providing inhibitory projections to other nuclei.

## III. Results

In order to evaluate the relationship between the DTI and transfer function measures, for each patient, we removed the outliers and kept the samples within 3 standard deviation from the mean. We removed the samples for which the max gain occurs below 1.5 Hz as the data could be distorted in vicinity of 1 Hz cutoff frequency. We employed linear mixed effect model (LME) to model the PF ratio and the maximum gain in relation to FA, AD, N, and L for group analysis. LME provides a robust framework for examining and assessing the strength and significance of the linear relationships between our predictors (FA, AD, N, and L) and our outcomes of interest (maximum gain and PF ratio). Each predictor variable was chosen based on theoretical considerations and previous empirical findings suggesting their relevance to the functional connectivity. One limitation of using linear models is the multicollinearity among features. Therefore, the Variance Inflation Factors (VIF) were calculated to ensure that collinearity is negligible in the models, affirming the robustness and reliability of our statistical results. After evaluating multicollinearity, we found that AD and FA are highly positively correlated (it can also be inferred from the FA formula presented in Equation 1). Therefore, in our analysis, we considered only FA; however, in our discussion, due to their positive correlation, we addressed both parameters. Their relationship is illustrated in Figure 3.

Figure 2 shows a visualization of three distinct pathways from the GPi to VoaVop in a single patient, exemplifying the individual fiber tracts and their corresponding PF ratio and maximum gains. The significance of the estimated coefficients were then evaluated by t-tes using using Satterthwaite’s method with the significance treshold of 0.05. We adjusted all the p-values using Bonferroni method after. All the statistical analysis were done in R-studio [34].

**Fig. 2:**
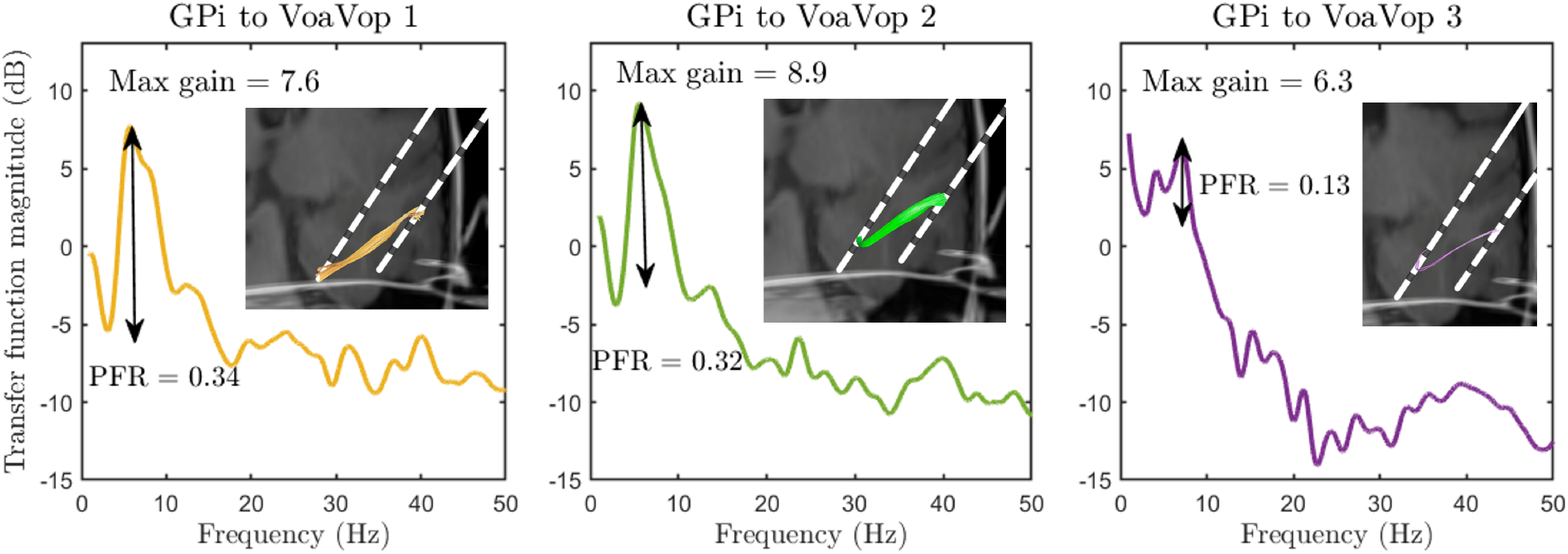
This figure illustrates a sample of three unique neural pathways from GPi to VoaVop for one patient, each characterized by varying FA, AD, length, and number of fibers. Accompanying each pathway is its respective transfer function magnitude plot. Maximum gain and the PF ratio are annotated on each plot for clarity. The larger fibers represented on the left (*FA* = 0.40, *AD* = 1.35, *L* = 16.5, *total N* = 3) and middle (*FA* = 0.42, *AD* = 1.38, *L* = 18.8, *total N* = 132) plots exhibit higher maximum gain and PF ratio values. Conversely, the pathway depicted on the right (*FA* = 0.38, *AD* = 1.2, *L* = 20.07, *total N* = 45) plot, characterized by smaller FA and AD has lower maximum gain and PF ratio, underscoring the relationship between the structural characteristics and functional connectivity.

**Fig. 3:**
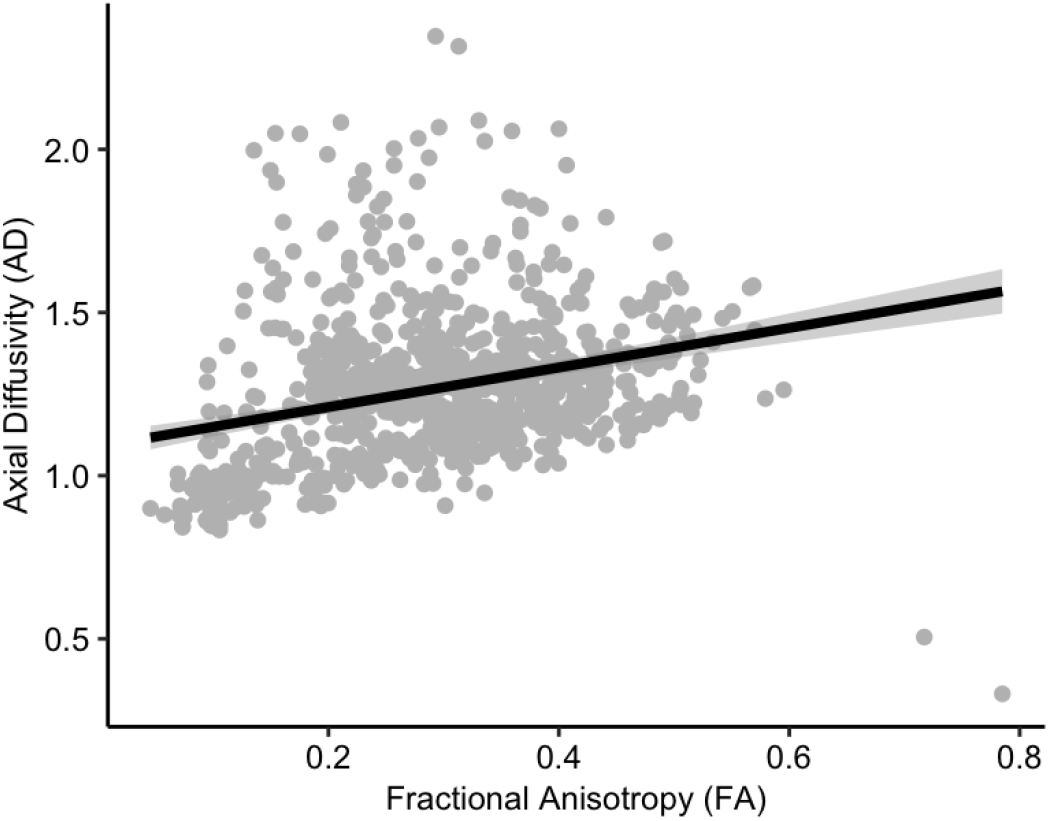
Relationship between the Fractional Anisotropy (FA) and Axial Diffusivity (AD) correlation. To avoid the issues caused my multicolinearity in linear mixed effect model we removed AD the formula and only considered FA.

Evaluation of the relationship between the PF ratio (*R*^2^ = 0.25) and maximum gain (*R*^2^ = 0.13) with FA, N, and L indicated a significant positive correlation between both the PF ratio and maximum gain with FA and perhaps AD, as illustrated in Fig. 4 and Fig. 5. Conversely, the analysis showed no statistically significant correlation between PF ratio and maximum gain with either fiber length (L) or the number of fibers per unit area (N) (Figures 4 and 5). The results of the LME analysis are presented in Table II, where we detail the estimated effects, confidence intervals, and statistical significance of each predictor.

**TABLE 2:** Statistical outcomes from the linear mixed effect models fit for PF ratio and maximum gain in relation to FA, AD, N, and L.

| DTI measures | Intercept [CI] | FA [CI] | N [CI] | L [CI] | $R^2$ |
| --- | --- | --- | --- | --- | --- |
| PF ratio | **34.63**<br>[32.13, 37.13] | **6.12**<br>[1.38, 10.86] | 1.44<br>[-1.10, 3.98] | -0.01<br>[-0.02, 0.00] | .25<br>(N=12) |
| Maximum Gain | **26.31**<br>[24.17, 28.44] | **6.77**<br>[1.78, 11.76] | 1.19<br>[-1.52, 3.89] | 0.0<br>[-0.02, 0.01] | .13<br>(N=12) |

**Fig. 4:**
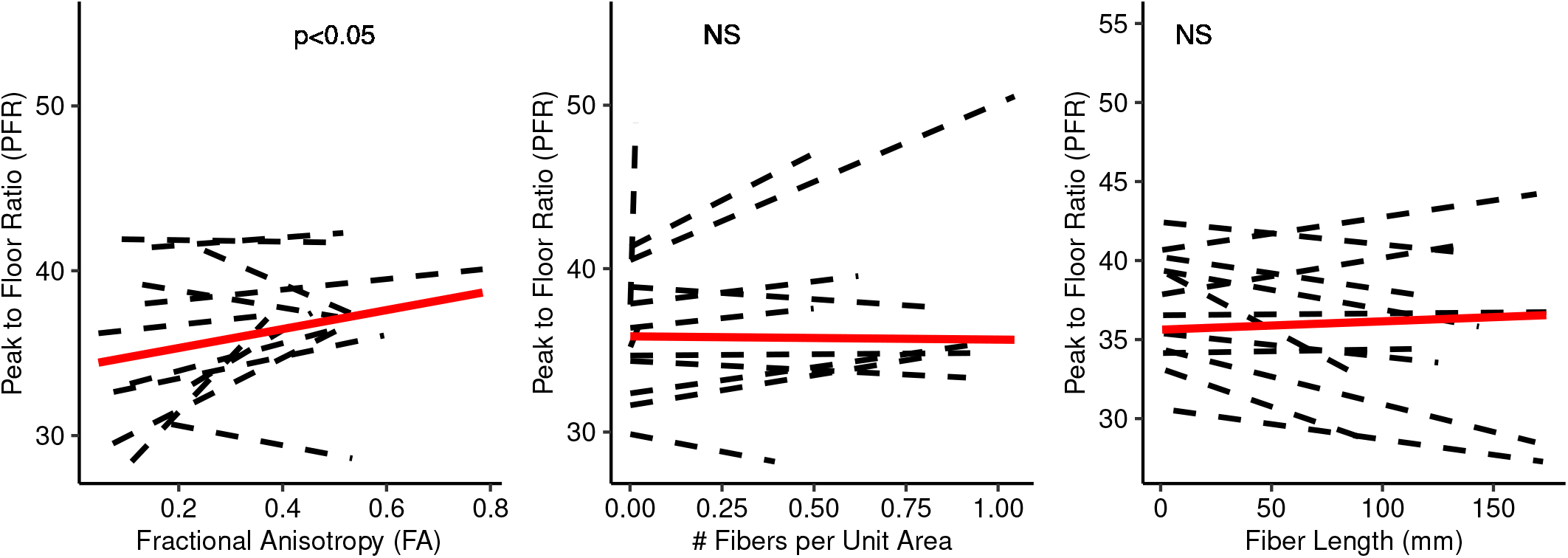
Illustration of the linear fits and the statistical significance between the PF ratio and DTI measures. Individual subject PF ratios are depicted by dashed black lines and the solid red lines represent the linear fit for group analysis. The figure highlights the significant correlation (*p −value <* 0.01) of PF ratio with Functional Anisotropy (FA) and the absence of significant correlation (NS) of PF ratio with number of tracts per unit area (N) and fiber length (L).

**Fig. 5:**
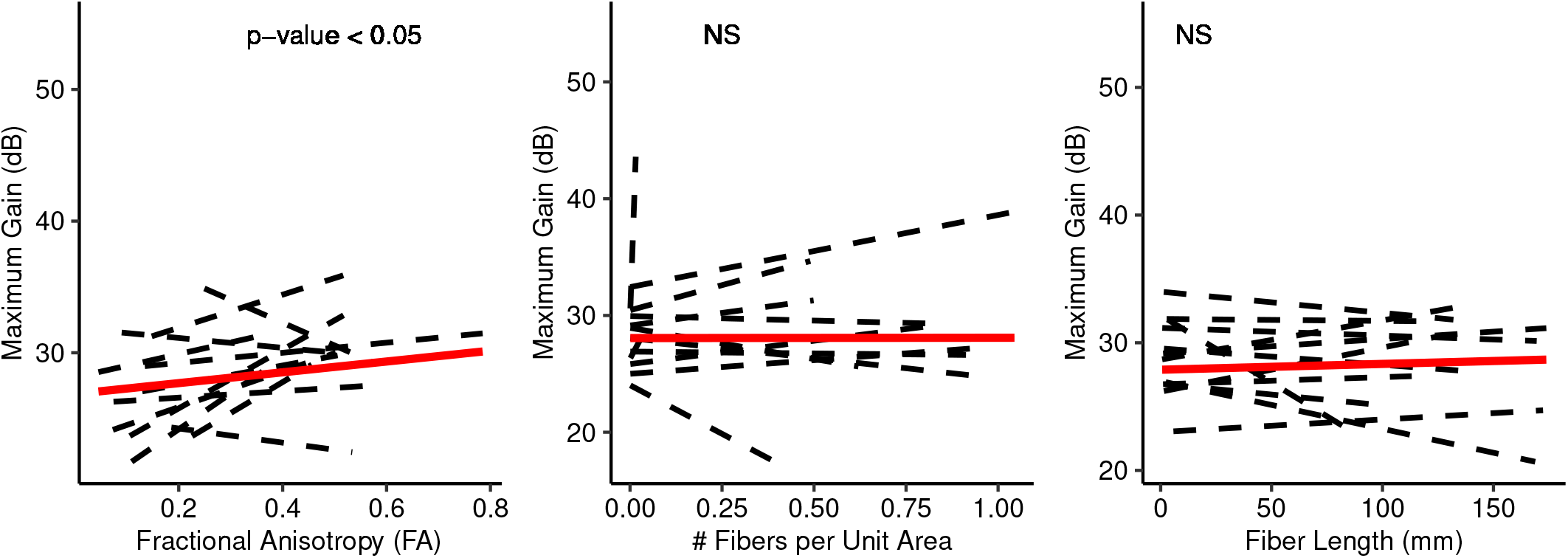
Illustration of the linear fits and the statistical significance between the maximum gains and DTI measures. Individual subject maximum gains are depicted by dashed black lines and the solid red lines represent the linear fit for group analysis. The figure highlights the significant correlation (*p− value <* 0.01) of maximum gain with Functional Anisotropy (FA) and the absence of a correlation of maximum gain with number of tracts per unit area (N) and fiber length (L).

## IV. Discussion

Several research studies to date focused on integrating structural and functional connectivity in order to understand how they correlate and the mutual information they share with one another [35]–[42]. These investigations predominantly depend on modalities such as electro-encephalography (EEG) and fMRI for assessing functional connectivity, while employing DTI to assess structural connectivity. However, despite significant insights offered by these studies, this relationship is still poorly understood. Notably, limitations such as the relatively low spatial resolution of EEG and the low temporal resolution of fMRI prevents us from conducting precise localization and capturing of rapid neural dynamics while assessing functional connectivity [43], [44]. Thus, it seems necessary to employ other modalities to fully address the relationship between structural and functional connectivity in the brain.

Here in this paper, we aimed to study the relationship between DTI parameters and characteristics of patient-specific transfer function models obtained from brain intrinsic neural activity. Thus, we investigated the correlation between FA, AD, Number of fiber tracts per unit area (N), and fiber tract length (L) as DTI parameters with maximum transfer function gain and PF ratio. Our results, consistent with previous works done on the relationship between the functional and anatomical connectivity [35]–[42], provide further evidence of the relationship between anatomical and functional connectivity. In particular, our results highlight positive correlation between FA and AD with both characteristic of transfer function models (i.e., PF ratio and maximum gain). Significant positive correlation between FA and maximum transfer function gain shows that axonal fibers with higher integrity can better amplify and spread an intrinsic brain signal throughout the brain network. FA significant positive correlation with PF ratio suggests that axonal fibers with higher integrity have the capacity to provide larger system response and signal transmission. Significant positive correlation of maximum transfer function gain and PF ratio with AD suggests that the maximum level of signal amplification and fast magnitude drop-offs happen with higher magnitudes of diffusion parallel to fiber tracts.

In addition, Abe et al. previously provided evidence on the correlation between DTI parameters (i.e., FA, tract length, and tract diameter) and characteristics of DBS evoked potentials (EPs) [29]. Their results suggest that the integrity of white matter tracts plays a crucial role in determining the efficiency, strength, and transmission speed of DBS-induced signals. Moreover, Kasiri et al. [21] demonstrated through a patient-specific transfer function approach that DBS pulses travel along normal pathways from the stimulation site to distant targets in the brain. These studies collectively highlight the varied role of DTI parameters, such as FA and AD, not only in providing insights into anatomical connectivity, but also in explaining information about neural signal transmission in the brain.

## V. Limitation

As a non-invasive imaging technique, DTI provides valuable information about the white matter microstructure which enables their visualization in 3D and facilitates studies on brain injures. However, similar to any imaging technique it has certain limitations. First, such computations of anatomical connectivity relies on the orientation of fibers which might result in inaccuracies in regions where the fibers interactions are complex [45]. Second, DTI has low image resolution, limiting its ability to identify single nerve fibers or small fiber bundles. Moreover, it is sensitive to noise and artifacts; therefore we require precise motion control and post-processing approaches. To better utilize DTI, integration with other imaging techniques and clinical data is necessary [45]. This is why we, previously, used DBS measurements to validate and confirm the effectiveness and accuracy of our DTI measurements for clinical applications [29].

In addition, it is crucial to acknowledge the limitations associated with Linear Time-Invariant (LTI) models, including transfer function analysis, which we employed to model the signal transmission between different nuclei in the deep brain regions. LTI models may not fully and accurately capture the dynamic and nonlinear nature of brain function. The brain’s complex and adaptive nature might involve time-varying dynamics that cannot be adequately addressed by LTI models.

Moreover, the transfer function gains here do not indicate that the information is transmitted through a direct pathway from input to the output of the pathway’s system. Such measure can also indicate a common input to both system’s input and output (two ends of a pathway), whether there is a fiber between them or not. However, in this study, since we are investigating the correlation of anatomical connectivity with the functional connectivity, there always exists a tract between the input and output. Therefore, this concern in not valid here, although it is a valid concern in general.

## VI. Conclusion

In conclusion, our study provides further evidence for the existence of significant relationship between structural and functional connectivity, offering valuable insights into how white matter integrity affects intrinsic neural activity propagation in the brain network.

## Data Availability

All data produced in the present study are available upon reasonable request to the authors

## Acknowledgements

We thank our volunteers and their parents for participating in this study. We also thank Jennifer MacLean, Diana Ferman, and Teresa Serna for their assistance with neurological examinations and Jaya Nataraj for helping in data collection. This study is funded by the Cerebral Palsy Alliance Research Foundation (PG02518).

## Conflicts of Interest

All authors declare no conflicts of interest.

## Authors Contributions

M.K., E.H.M., R.S., S.A.S.M., and T.D.S. conducted the experiments and collected the data. M.K. analyzed the neurophysiological data and performed statistical analysis. S.A. performed the image analysis and electrode localization. R.S. integrated DTI and neurophysiology data. M.K., S.A., and T.D.S. interpreted the results of the study. T.D.S. performed the neurological examination. All authors contributed to writing and revision of the final manuscript.

## Notes

### Competing Interest Statement

The authors have declared no competing interest.

### Author Declarations

Institutional review board of the Children's Health Orange County (CHOC) hospital gave ethical approval for this work. Institutional review board Children's Hospital Los Angeles (CHLA) gave ethical approval for this work.

### Summary of Updates

We improved the analysis. They results are the same.

## References

[1] C. Yen, C.-L. Lin, and M.-C. Chiang, “Exploring the frontiers of neuroimaging: a review of recent advances in understanding brain functioning and disorders,” Life, vol. 13, no. 7, p. 1472, 2023.

[2] L. Kong, Q. He, Q. Li, R. Schreiber, K. I. Kaitin, and L. Shao, “Rapid progress in neuroimaging technologies fuels central nervous system translational medicine,” Drug Discovery Today, p. 103485, 2023.

[3] J. M. Henderson, ““connectomic surgery”: diffusion tensor imaging (dti) tractography as a targeting modality for surgical modulation of neural networks,” Frontiers in integrative neuroscience, vol. 6, p. 15, 2012.

[4] V. A. Coenen and M. Reisert, “Dti for brain targeting: Diffusion weighted imaging fiber tractography—assisted deep brain stimulation,” in International Review of Neurobiology. Elsevier, 2021, vol. 159, pp. 47–67.

[5] S. Owen, J. Heath, M. Kringelbach, A. Green, E. Pereira, N. Jenkinson, T. Jegan, J. Stein, and T. Aziz, “Pre-operative dti and probabilisitic tractography in four patients with deep brain stimulation for chronic pain,” Journal of Clinical Neuroscience, vol. 15, no. 7, pp. 801–805, 2008.

[6] B. Mädler and V. Coenen, “Explaining clinical effects of deep brain stimulation through simplified target-specific modeling of the volume of activated tissue,” American Journal of Neuroradiology, vol. 33, no. 6, pp. 1072–1080, 2012.

[7] V. A. Coenen, N. Allert, S. Paus, M. Kronenbürger, H. Urbach, and B. Mädler, “Modulation of the cerebello-thalamo-cortical network in thalamic deep brain stimulation for tremor: a diffusion tensor imaging study,” Neurosurgery, vol. 75, no. 6, pp. 657–670, 2014.

[8] K.N. Aaq, “Improving surgical outcome using diffusion tensor imaging techniques in deep brain stimulation. front surg. 2017; 4: 1–11,” 2017.

[9] S. Owen, J. Heath, M. Kringelbach, J. Stein, and T. Aziz, “Preoperative dti and probabilistic tractography in an amputee with deep brain stimulation for lower limb stump pain,” British journal of neurosurgery, vol. 21, no. 5, pp. 485–490, 2007.

[10] V. A. Coenen, B. Mädler, H. Schiffbauer, H. Urbach, and N. Allert, “Individual fiber anatomy of the subthalamic region revealed with diffusion tensor imaging: a concept to identify the deep brain stimulation target for tremor suppression,” Neurosurgery, vol. 68, no. 4, pp. 1069–1076, 2011.

[11] J. M. Anthofer, K. Steib, C. Fellner, M. Lange, A. Brawanski, and J. Schlaier, “Dti-based deterministic fibre tracking of the medial forebrain bundle,” Acta neurochirurgica, vol. 157, pp. 469–477, 2015.

[12] V. A. Coenen, B. Varkuti, Y. Parpaley, S. Skodda, T. Prokop, H. Urbach, M. Li, and P. C. Reinacher, “Postoperative neuroimaging analysis of drt deep brain stimulation revision surgery for complicated essential tremor,” Acta Neurochirurgica, vol. 159, pp. 779–787, 2017.

[13] E. W. Lang, A.M. Tomé, I. R. Keck, J. Górriz-Sáez, and C. G. Puntonet, “Brain connectivity analysis: a short survey,” Computational intelligence and neuroscience, vol. 2012, pp. 8–8, 2012.

[14] J. B. Rowe, “Connectivity analysis is essential to understand neurological disorders,” Frontiers in Systems Neuroscience, vol. 4, p. 144, 2010.

[15] P. M. Rossini, R. Di Iorio, M. Bentivoglio, G. Bertini, F. Ferreri, C. Gerloff, R. J. Ilmoniemi, F. Miraglia, M. A. Nitsche, F. Pestilli et al., “Methods for analysis of brain connectivity: An ifcn-sponsored review,” Clinical Neurophysiology, vol. 130, no. 10, pp. 1833–1858, 2019.

[16] S. J. Carr, A. Gershon, N. Shafiabadi, S. D. Lhatoo, C. Tatsuoka, and S. S. Sahoo, “An integrative approach to study structural and functional network connectivity in epilepsy using imaging and signal data,” Frontiers in integrative neuroscience, vol. 14, p. 491403, 2021.

[17] J. Goñi, M. P. Van Den Heuvel, A. Avena-Koenigsberger, N. Velez de Mendizabal, R. F. Betzel, A. Griffa, P. Hagmann, B. Corominas-Murtra, J.-P. Thiran, and O. Sporns, “Resting-brain functional connectivity predicted by analytic measures of network communication,” Proceedings of the National Academy of Sciences, vol. 111, no. 2, pp. 833–838, 2014.

[18] M. D. Greicius, K. Supekar, V. Menon, and R. F. Dougherty, “Resting-state functional connectivity reflects structural connectivity in the default mode network,” Cerebral cortex, vol. 19, no. 1, pp. 72–78, 2009.

[19] M. J. Rosa, J. Kilner, F. Blankenburg, O. Josephs, and W. Penny, “Estimating the transfer function from neuronal activity to bold using simultaneous eeg-fmri,” Neuroimage, vol. 49, no. 2, pp. 1496–1509, 2010.

[20] G. Kamali, R. J. Smith, M. Hays, C. Coogan, N. E. Crone, J. Y. Kang, and S. V. Sarma, “Transfer function models for the localization of seizure onset zone from cortico-cortical evoked potentials,” Frontiers in Neurology, vol. 11, p. 579961, 2020.

[21] M. Kasiri, J. W. Hillman, E. Hernandez-Martin, S. A. S. Mousavi, and T. D. Sanger, “Endogenous signals during active movement predict deep brain stimulation evoked potential pathways: Results of a transfer function analysis,” medRxiv, pp. 2023–04, 2023.

[22] M. Liker and T. Sanger, “Pediatric deep brain stimulation in secondary dystonia using stereotactic depth electrode targeting,” in MOVEMENT DISORDERS, vol. 34. WILEY 111 RIVER ST, HOBOKEN 07030-5774, NJ USA, 2019, pp. S534–S535.

[23] M. A. Liker, T. D. Sanger, J. A. MacLean, J. Nataraj, E. Arguelles, M. Krieger, A. Robison, and J. Olaya, “Stereotactic awake basal ganglia electrophysiological recording and stimulation (sabers): A novel staged procedure for personalized targeting of deep brain stimulation in pediatric movement and neuropsychiatric disorders,” Journal of Child Neurology, vol. 0, no. 0, p. 08830738231224057, 0, pMID: 38409793. [Online]. Available: 10.1177/08830738231224057

[24] N. e. a. Boy, “Proposed recommendations for diagnosing and managing individuals with glutaric aciduria type i: second revision,” Journal of inherited metabolic disease, vol. 40, no. 1, pp. 75–101, 2017.

[25] L. Abela and M. A. Kurian, “Kmt2b-related dystonia synonyms: Dyt28, dyt-kmt2b,” 1993. [Online]. Available: https://www.ncbi.nlm.nih.gov/books/

[26] T. D. Sanger, “Deep brain stimulation for cerebral palsy: where are we now?” Developmental Medicine & Child Neurology, vol. 62, no. 1, pp. 28–33, 2020. [Online]. Available: https://onlinelibrary.wiley.com/doi/abs/10.1111/dmcn.14295

[27] E. Hernandez-Martin, M. Kasiri, S. Abe, J. MacLean, J. Olaya, M. Liker, J. Chu, and T. D. Sanger, “Globus pallidus internus activity increases during voluntary movement in children with dystonia,” iScience, vol. 26, no. 7, p. 107066, 2023. [Online]. Available: https://www.sciencedirect.com/science/article/pii/S2589004223011434

[28] P. A. Taylor, A. Alhamud, A. van der Kouwe, M. G. Saleh, B. Laughton, and E. Meintjes, “Assessing the performance of different dti motion correction strategies in the presence of epi distortion correction,” Human brain mapping, vol. 37, no. 12, pp. 4405–4424, Dec 2016. [Online]. Available: 10.1002/hbm.23318

[29] S. Abe, J. Vidmark, E. Hernandez-Martin, M. Kasiri, R. Sorouhmojdehi, S. A. S. Mousavi, and T. D. Sanger, “Diffusion tractography predicts deep brain stimulation evoked potential amplitude and delay,” medRxiv, 2024. [Online]. Available: https://www.medrxiv.org/content/early/2024/04/13/2024.04.11.24305627

[30] P. J. Winklewski, A. Sabisz, P. Naumczyk, K. Jodzio, E. Szurowska, and A. Szarmach, “Understanding the physiopathology behind axial and radial diffusivity changes—what do we know?” Frontiers in Neurology, vol. 9, 2018. [Online]. Available: https://www.frontiersin.org/journals/neurology/articles/10.3389/fneur.2018.00092

[31] “Chapter 7 - new image contrasts from diffusion tensor imaging: Theory, meaning, and usefulness of dti-based image contrast,” in Introduction to Diffusion Tensor Imaging (Second Edition), second edition ed., S. Mori and J.-D. Tournier, Eds. San Diego: Academic Press, 2014, pp. 53–64. [Online]. Available: https://www.sciencedirect.com/science/article/pii/B9780123983985000072

[32] D. K. Jones, T. R. Knösche, and R. Turner, “White matter integrity, fiber count, and other fallacies: The do’s and don’ts of diffusion mri,” NeuroImage, vol. 73, pp. 239–254, 2013. [Online]. Available: https://www.sciencedirect.com/science/article/pii/S1053811912007306

[33] S. Correia, S. Y. Lee, T. Voorn, D. F. Tate, R. H. Paul, S. Zhang, S. P. Salloway, P. F. Malloy, and D. H. Laidlaw, “Quantitative tractography metrics of white matter integrity in diffusion-tensor mri,” NeuroImage, vol. 42, no. 2, pp. 568–581, 2008. [Online]. Available: https://www.sciencedirect.com/science/article/pii/S1053811908006435

[34] D. Bates, M. Mächler, B. Bolker, and S. Walker, “Fitting linear mixed-effects models using lme4,” Journal of Statistical Software, vol. 67, pp. 1–48, 2015. [Online]. Available: https://www.jstatsoft.org/index.php/jss/article/view/v067i01

[35] M. C. Litwińczuk, N. Muhlert, L. Cloutman, N. Trujillo-Barreto, and A. Woollams, “Combination of structural and functional connectivity explains unique variation in specific domains of cognitive function,” NeuroImage, vol. 262, p. 119531, 2022.

[36] P. Babaeeghazvini, L. M. Rueda-Delgado, J. Gooijers, S. P. Swinnen, and A. Daffertshofer, “Brain structural and functional connectivity: A review of combined works of diffusion magnetic resonance imaging and electro-encephalography,” Frontiers in Human Neuroscience, vol. 15, p. 721206, 2021.

[37] E. Gleichgerrcht, A. S. Greenblatt, T. S. Kellermann, N. Rowland, W. A. Vandergrift, J. Edwards, K. A. Davis, and L. Bonilha, “Patterns of seizure spread in temporal lobe epilepsy are associated with distinct white matter tracts,” Epilepsy research, vol. 171, p. 106571, 2021.

[38] T. L. Richards, T. J. Grabowski, P. Boord, K. Yagle, M. Askren, Z. Mestre, P. Robinson, O. Welker, D. Gulliford, W. Nagy et al., “Contrasting brain patterns of writing-related dti parameters, fmri connectivity, and dti–fmri connectivity correlations in children with and without dysgraphia or dyslexia,” NeuroImage: Clinical, vol. 8, pp. 408–421, 2015.

[39] H. Huang and M. Ding, “Linking functional connectivity and structural connectivity quantitatively: a comparison of methods,” Brain connectivity, vol. 6, no. 2, pp. 99–108, 2016.

[40] R. Schmidt, E. Verstraete, M. A. de Reus, J. H. Veldink, L. H. van den Berg, and M. P. van den Heuvel, “Correlation between structural and functional connectivity impairment in amyotrophic lateral sclerosis,” Human brain mapping, vol. 35, no. 9, pp. 4386–4395, 2014.

[41] M. A. Koch, D. G. Norris, and M. Hund-Georgiadis, “An investigation of functional and anatomical connectivity using magnetic resonance imaging,” Neuroimage, vol. 16, no. 1, pp. 241–250, 2002.

[42] H. Liu, G. Fan, K. Xu, and F. Wang, “Changes in cerebellar functional connectivity and anatomical connectivity in schizophrenia: a combined resting-state functional mri and diffusion tensor imaging study,” Journal of magnetic resonance imaging, vol. 34, no. 6, pp. 1430–1438, 2011.

[43] G. H. Glover, “Overview of functional magnetic resonance imaging,” Neurosurgery Clinics, vol. 22, no. 2, pp. 133–139, 2011.

[44] B. Burle, L. Spieser, C. Roger, L. Casini, T. Hasbroucq, and F. Vidal, “Spatial and temporal resolutions of eeg: Is it really black and white? a scalp current density view,” International Journal of Psychophysiology, vol. 97, no. 3, pp. 210–220, 2015.

[45] D. K. Jones, “Challenges and limitations of quantifying brain connectivity in vivo with diffusion mri,” Imaging in Medicine, vol. 2, pp. 341–355, 2010. [Online]. Available: https://api.semanticscholar.org/CorpusID:73108365

